# High prevalence of *Plasmodium malariae* and *Plasmodium ovale* in co-infections with *Plasmodium falciparum* in asymptomatic malaria parasite carriers in Southwest Nigeria

**DOI:** 10.1101/2021.01.14.21249635

**Authors:** Muhydeen Abiodun Abdulraheem, Medard Ernest, Ifeoma Ugwuanyi, Hussein M. Abkallo, Saori Nishikawa, Mofeyisade Adeleke, Adebola E. Orimadegun, Richard Culleton

**Affiliations:** Malaria Unit, Department of Pathology, Institute of Tropical Medicine, Nagasaki University, 1-12-4 Sakamoto, Nagasaki 852-8523, Japan; Diana Princess of Wales Hospital, Grimsby, Northeast Lincolnshire, UK; Laboratory of Malaria and Vector Research, National Institute of Allergy and Infectious Diseases, National Institutes of Health, Rockville, MD 20852, USA; University of New South Wales (UNSW), Sydney, Australia; International Livestock Research Institute (ILRI), P.O. Box 30709 Nairobi 00100 Kenya; Graduate School of Social and Cultural Science, Kumamoto University, Japan; Moniya General Hospital, Akinyele Local Government, Nigeria; Institute of Child Health College of Medicine University of Ibadan, Nigeria; Department of Protozoology, Institute of Tropical Medicine, Nagasaki University, 1-12-4 Sakamoto, Nagasaki 852-8523, Japan; Division of Molecular Parasitology, Proteo-Science Centre, Ehime University, Shitsukawa, Toon, Ehime 791-0295, Japan

**Keywords:** *Plasmodium falciparum*, *Plasmodium malariae*, *Plasmodium ovale*, asymptomatic malaria, Nigeria

## Abstract

Asymptomatic malaria parasite carriers do not seek anti-malarial treatment and may constitute a silent infectious reservoir. In order to assess the level of asymptomatic and symptomatic carriage amongst adolescents in a highly endemic area, and to identify the risk factors associated with such carriage, we conducted a cross sectional survey of 1032 adolescents (ages 10-19) from eight schools located in Ibadan, Southwest Nigeria in 2016. Blood films and blood spot filter paper samples were prepared for microscopy and DNA analysis. The prevalence of asymptomatic malaria was determined using microscopy, rapid diagnostic tests and PCR for 658 randomly selected samples. Of these, we found that 80% of asymptomatic schoolchildren were positive for malaria parasites by PCR, compared to 47% and 9% determined by RDT and microscopy, respectively. Malaria parasite species typing was performed using PCR targeting the mitochondrial *CoxIII* gene, and revealed high rates of carriage of *Plasmodium malariae* (53%) and *Plasmodium ovale* (24%). Most asymptomatic infections were co-infections of two or more species (62%), with *P. falciparum* + *P. malariae* the most common (35%), followed by *P. falciparum* + *P. malariae* + *P. ovale* (21%) and *P. falciparum* + *P. ovale* (6%). Single infections of *P. falciparum, P. malariae* and *P. ovale* accounted for 24%, 10% and 4% of all asymptomatic infections respectively. To compare the species composition of asymptomatic and symptomatic infections, further sample collection was carried out in 2017 at one of the previously sampled schools, and at a nearby hospital. Whilst the species composition of the asymptomatic infections was similar to that observed in 2016, the symptomatic infections were markedly different, with single infections of *P. falciparum* observed in 91% of patients, *P. falciparum* + *P. malariae* in 5% and *P. falciparum* + *P. ovale* in 4%.

## 1. Introduction

In areas of high malaria parasite transmission, a proportion of the population will carry malaria parasites but not display the clinical symptoms of malaria. Such asymptomatic malaria carriage is thought to result from the early acquisition of immunity achieved through frequent exposure to the parasite. This exposure enables the immune response to limit parasite growth to low parasitaemias, whilst also avoiding the development of the inflammatory immune responses that characterise the typical clinical manifestations of the disease (Galatas et al., 2016). Anti-disease immunity (or tolerance) allows the carriage of parasites without the development of symptoms, and this, combined with anti-parasite immunity that suppresses parasite loads in previously infected individuals leads to the establishment of low-parasitaemia, chronic asymptomatic malaria parasite carriage (Daubersies et al., 1996; Day and Marsh, 1991; Trape et al., 1994). The parasitaemias of these infections are typically below the threshold of detection by microscopy, but may be detected by more sensitive diagnostics such as PCR.

Asymptomatic malaria infections often go undetected and untreated, and can result in a hidden reservoir for transmission (Alves et al., 2005). This poses challenges for the control and elimination of malaria in high transmission regions such as the south-west of Nigeria. For example, a study in Senegal suggested that more than 90% of exposed individuals were likely infected with chronic asymptomatic malaria (Bottius et al., 1996).

A number of studies have reported asymptomatic malaria in Nigeria (Achidi et al., 1995; Anorlu et al., 2001; Jeremiah and Uko, 2007; Kotila et al., 2007; Nwagha et al., 2011; Nwagha et al., 2009; Nwaneri et al., 2013; Ojurongbe et al., 2011). However, these studies are mostly descriptive and extensive analytical studies are lacking. Most reports are limited to general prevalence surveys, without extensive molecular analyses and quantification of parasite burdens. Identifying and targeting these infections is important for malaria control, as they constitute significant infectious reservoirs for the continued transmission of the parasite.

Despite increased resources for malaria control, including initiatives such as Roll Back Malaria, the Multilateral Initiative on Malaria and the introduction of artemisinin-based combination therapy, malaria remains a major threat to the health and lives of children in Nigeria. Indeed, Nigeria accounts for 25% of the malaria burden of the entire African continent (Organization, 2019).

The vast majority of malariometric studies conducted in Nigeria, and in Sub-Sharan Africa in general, have focussed on the parasite burden in young children, or in adults. Generally, there is a dearth of data on the malaria burden among adolescents (aged 10-19) in sub-Saharan Africa. Furthermore, the current malaria control measures and strategies focus on children under five years old and pregnant women, with adolescents remaining neglected. There is no control programme that offers guidelines on the use of measures such as insecticide treated nets, indoor residual spraying and chemoprophylaxis for the control of malaria in this age group (FMOH, 2008). Yet, these adolescents, especially those in schools, constitute a considerable proportion of the at-risk population.

We hypothesised that schooling adolescents in high transmission areas such as southwest Nigeria might constitute a significant infectious reservoir of malaria parasites due to the potentially high percentage of asymptomatic malaria parasite carriers amongst this demographic. In order to better understand the dynamics of malaria parasite carriage amongst this age group, we carried out a cross-sectional survey of schooling adolescents in Ibadan, southwest Nigeria, involving blood sampling for malaria parasite diagnosis, and the use of an interviewer-administered questionnaire to identify possible risk factors for parasite carriage. We recruited 1032 adolescents with no malaria symptoms from a total of eight secondary schools in the region, and collected blood samples for microscopy, RDT and PCR diagnosis.

As previous surveys of the prevalence of non-falciparum malaria parasites have been conducted on symptomatic infections, we also assayed the prevalence of *P. malariae* and *P. ovale* in asymptomatic parasite carriers by PCR.

## 2. Methods

### 2.1. Study design, setting and population

A cross-sectional study involving schooling adolescents aged 10 to 19 years living in two Local Government Areas (LGAs) in Ibadan (Ibadan North and Akinyele), Oyo State, Nigeria was carried out **(Figure 1)**. The choice of the study settings and population for this study were based on the fact that substantial parts of the Akinyele area has rural communities while Ibadan North is essentially urban. The population distributions of the two LGAs are similar. According to the 2006 population census, individuals aged 10-19 years constitute about 5% of the 110,000 estimated population of the two LGAs. There are about 43 officially registered public secondary schools in the two LGAs.

**Figure 1.**
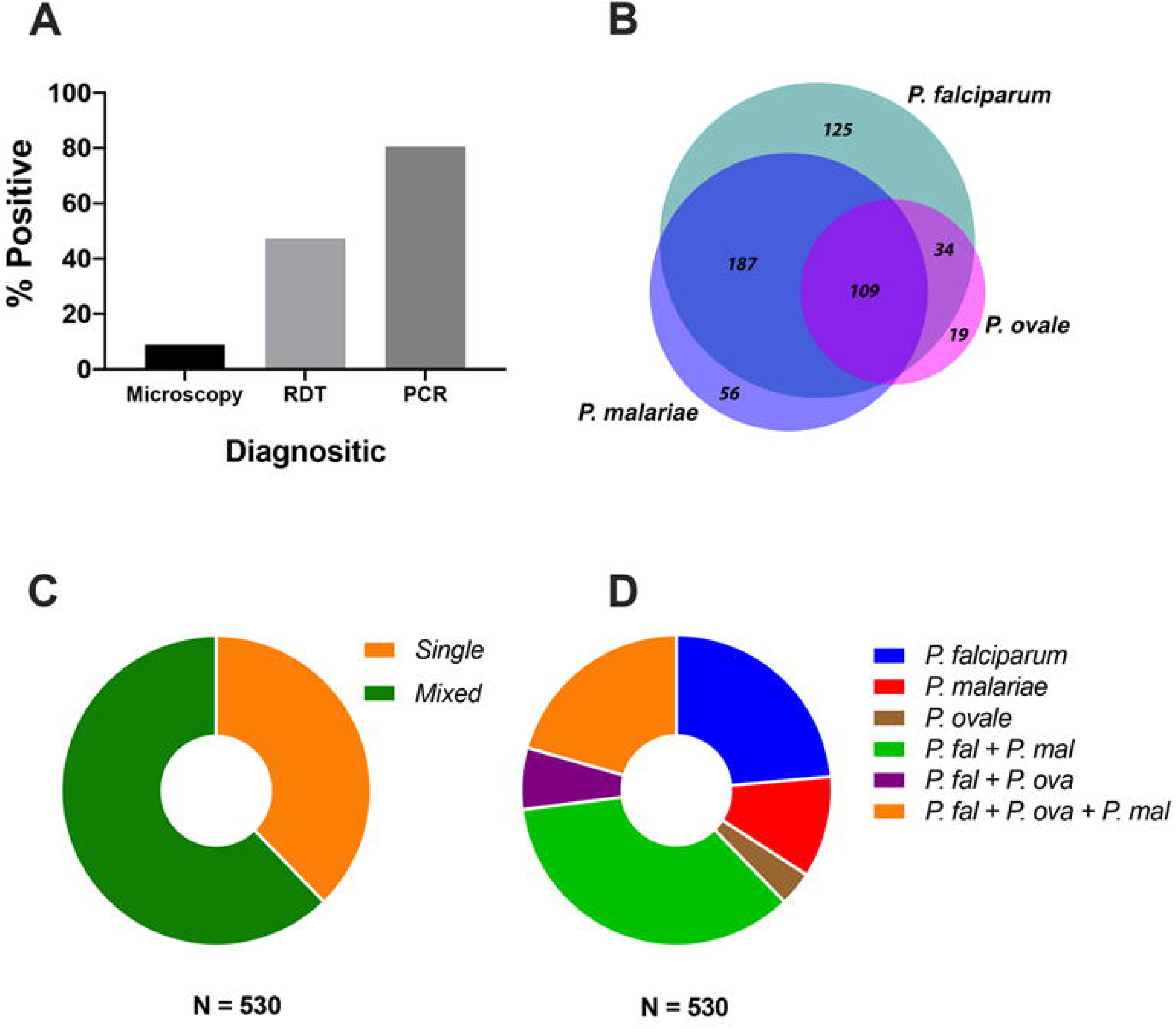
Species composition of 658 asymptomatic malaria parasite infections. (A) Percentage positivity rate for microscopy, rapid diagnostic test and PCR for 658 randomly selected samples from a total of 1032 blood samples collected from schooling adolescents (aged 10-19) in Ibadan province, Southwest Nigeria. (B) Prevalence of each of three malaria parasite species assayed (*Plasmodium falciparum, Plasmodium malariae* and *Plasmodium ovale*). (C) The percentage of single and mixed species infections. (D) Malaria parasite species composition of parasite infected blood samples.

### 2.2. Sample size determination

We used the one-sample comparison of proportion in StatCalc Menu of Epi Info 7.0.9.34 (CDC, 2012) to estimate the required sample size. We assumed that the prevalence of *P. falciparum, P. malariae* and *P. ovale* were 46.6, 12.9 and 8.3 %, respectively obtained in a recent study in Democratic Republic of the Congo (DRC) (Doctor et al., 2016) which has a similar malaria burden to that of Nigeria. A minimum of 197 participants was required to achieve 90% power at 95% level of confidence when the allowable margin of error of 5% and prevalence of 8.3% were used for the estimation. However, considering the clustering nature of our sampling and non-responses, we adopted a design effect factor of 3 and non-response rate of 10% and these increased the minimum sample size to 624.

### 2.3. Sampling of participants

A two-stage random sampling technique was employed to select eight secondary schools in the Ibadan North and Akinyele Local Government Area. A sampling frame of all potential recruitment sites was developed. The two Local Government Areas were already officially mapped into different geo-political wards by the local government councils and a school was randomly selected from each geo-political ward. If the number of adolescents recruited from any school was inadequate, then the next school on the sampling frame was chosen. Another sampling frame for all eligible children was made for each study centre. At stage two, adolescents were randomly selected from the list based on the ratio of the population of each selected school to overall population of all schools. We screened a total of 1032 adolescents for malaria rapid diagnostic test (mRDT) and 658 samples were randomly selected for molecular analysis. Of the 658 samples, there were 309 samples mRDT positive and 349 mRDT negative.

To recruit symptomatic participants, patients aged 10 to 25 years presenting with symptoms of malaria at the outpatient department of General Hospital Moniya, a secondary health care facility located within Akinyele local govt were enrolled in this study. The hospital provides various health care services to its immediate environs as well as neighbouring communities, it is mainly the first point of call for most patients seeking health care around Akinyele Local Government.

The inclusion criteria included consecutive patients with fever, malaise, prostration, nausea and vomiting who gave their consent through interviewer administered questionnaires and tested positive for malaria using RDT. Patients who did not give their consent to participate in the study and those who had used anti malaria before presentation were excluded.

### 2.4. Enrolment and Sample Collection from symptomatic malaria patients

Patients who gave their consent had their vital signs (temperature) and anthropometric measurements taken. Consent was given via either signature or thumb print. While maintaining asepsis, the pulp of the finger for needle prick was cleaned with an alcohol swab. Fingers were pricked using lancets and two drops of blood were dropped onto RDT kits and read after 15 minutes. If RDT positive, patients were informed and asked for consent to draw about two millilitres of blood for thick and thin film preparation, haemoglobin electrophoresis and DBS for PCR. All patients gave their consent. The site for sample collection was cleaned with an alcohol swab, a tourniquet was applied and with the aid of 26G needle and syringe about two millilitres of blood was drawn and transferred to an EDTA bottle. Pressure was applied at the site of collection to arrest haemorrhage. Samples were transported to the lab scientist for analysis on a daily basis.

### 2.5. Data collection methods

We made two visits to the selected schools. The first visit involved the selection of the pupils and the distribution of the information leaflets and consent forms to students for their parents/caregivers. The second visit involved the filling of the questionnaires, anthropometric measurements and collection of blood samples via skin prick. A validated structured interviewer-administered questionnaire was used to collect information on socio-demographic characteristics including age, sex, level of educational attainment and occupation of the parents, family living conditions, past medical history, and recent history of fever. The age of each student was verified from school/hospital records and/or birth certificates. Height and weight were measured according to World Health Organization (WHO) standards. Weight was measured using a battery powered digital scale (Seca, Inc, Columbia, MD, USA). Blood collection was carried out in a ‘cubicle’ or a room in each school. Samples and data were collected between November and December 2014. From the same skin-prick spot, samples for mRDT were prepared and few drops of blood were spotted on the two circles of Whatman filter paper for DNA analyses. The filter paper sample, after drying, was stored individually in a separate plastic bag, sealed with a desiccant at room temperature in Nigeria before transfer to Malaria Unit, Institute of Tropical Medicine, Nagasaki University, Japan.

### 2.6. DNA Extraction

DNA was extracted using the method described by Boom *et al*, ^108^ a guanidine based technique for extracting nucleic acid using prepared buffer solutions. DNA was extracted from a randomly selected 658 dried blood spots; due to resource and time restrictions, we were unable to process all 1032 samples.

### 2.7. Species Diagnosis PCR

The presence of malaria parasite DNA and diagnosis of parasite species was performed using a PCR based on the *cox3* gene, as previously described (Isozumi et al., 2015), with some modifications. Briefly, for the outer round of a nested PCR, 5 μl of DNA solution was added to a reaction containing 12.5 μl OneTaq^®^ MasterMix (New England Biolabs, USA), 0.5 μl of each primer (*MtUF* 5’-CTCGCCATTTGATAGCGGTTAACC-3’, and *MtUR* 5’-CCTGTTATCCCCGGCGAACCTTC-3’) at an initial concentration of 10 μM, and 6.5 μl of deionized water (total reaction volume, 25 μl). Thermocycler conditions were as follows; initial denaturation at 94°C for 30 seconds, followed by 40 cycles of 94°C for 30 seconds, 63°C for 60 seconds, 68°C for 60 seconds, followed by a final extension step of 68°C for 5 minutes. The inner round of the nested PCR was carried out using 2 μl of a 1:100 dilution of the PCR product from the outer round, added to a reaction containing 12.5 μl OneTaq^®^ MasterMix (New England Biolabs, USA), 0.5 μl of each primer (*P. falciparum* specific; *MtNst_falF* 5’-GAACACAATTGTCTATTCGTACAATTATTC-3’, and *MtNst_falR* 5’-GAACACAATTGTCTATTCGTACAATTATTC-3’, *P. malariae* specific; *MtNst_malF* 5’-CTAGCTTTGTACACAAATTAATTCGTCTAC-3’, and *MtNst_malR* 5’-CTTTATAAGAATGATAGATATTTATGACATA-3’, *P. ovale* specific; *MtNst_ov2F* 5’-ATTATTGTCAAATATAAGTACTTTAATC-3’ and *MtNst_ov2R* 5’-GGTTGAAGTTTATGATACTAATAAC-3’) at an initial concentration of 10 μM, and 9.5 μl of deionized water (total reaction volume, 25 μl). Thermocycler conditions for the inner round were as follows; initial denaturation at 94°C for 30 seconds, followed by 35 cycles of 94°C for 30 seconds, 56°C for 60 seconds, 68°C for 60 seconds, followed by a final extension step of 68°C for 5 minutes. PCR products were stained with ethidium bromide solution and visualised under UV light on 2% agarose gels run at 100 volts for 15 minutes.

### 2.8. Determination of the multiplicity of infection through PCR of the MSP1 locus

Determination of *P. falciparum* msp1 haplotypes was performed using nested PCR. We first amplified block 2/6 of the msp1 locus with primers flanking the conserved outer region, using the following conditions. 10 μl of OneTaq^®^ MasterMix (New England Biolabs, USA) 1 μl of each primer (Pfmsp1F: 5’-GCTTTAGAAGATGCAGTATTG-3’ and Pfmsp1R: 5’-CTTAAAGAATAATCTTCCATGATAT-3’) at initial concentration of 5 μM, 5 μl of DNA and 3 ul of deionized water to make 20 μl total reaction volume. Thermocycler conditions were as follows; initial denaturation at 94°C for 2 minutes, followed by 35 cycles at 94°C for 30 seconds, 42°C for 30 seconds, 68°C for 110 seconds followed by a final extension of 68°C for 5 minutes. PCR products were diluted 1000X for subsequent PCR reactions.

PCR amplification of block 2,4, and 6 was performed as previously described (Isozumi et al., 2015) with some modifications. Briefly, for PCR typing of block 2/6, 1 μl of DNA (from the 1000X diluted PCR products above) was added to a reaction containing 10μl OneTaq^®^ MasterMix, 1μl of each primer (*K2F*: 5’-TCTTAAATGAAGAAGAAATTACTACAAA-3’ or *M2F*: 5’-GGTTCAGGTAATTCAAGACGTAC-3’ or R2F: 5’-TAAAGGATGGAGCAAATACTCAAGT-3’ and K6R: 5’-GAATCAACTGAACCCAATGAATATC-3’ or *M6R*: 5’-CAAGATCCTACGAAATCTGTTCAAAT-3’) at initial concentration of 5 μM, 7 μl of deionized water to make 20 μl of total reaction volume. Thermocycler conditions were as follows; initial denaturation at 94C for 2 minutes, followed by 30 cycles of 94°C for 30 seconds, 52°C for 30 seconds, 68°C for 90 seconds, followed by a final extension step of 68°C for 5 minutes. 10 μl of PCR products were used for agarose (1%) gel electrophoresis, at 100 volts for 25-30 minutes. PCR products on agarose gel were stained with ethidium bromide solution and visualized under a UV light transilluminator.

For typing of block 4, 1 μl of 1000X diluted positive PCR products only (from block 2/6 typing) was added to a reaction containing 10 μl OneTaq^®^ MasterMix (New England Biolabs, USA), 1 μl of each primer (*K4aF*: 5’-AATGAAATTAAAAATCCCCCACCGG-3’ or M4aF: 5’-TTGAAGATATAGATAAAATTAAAACAGATG-3’ and *K4bR*: 5’-CTTGATAAGAACAAAAAAATCGAGGA-3’ or *M4bR*: 5’-CTGAGAATAAGAAAAAAGAAGTCGA-3’) at initial concentration of 5 μM and 7 μl of deionized water (Total reaction volume 20 μl). Thermocycler conditions were as follows; initial denaturation at 94°C for 2 minutes followed by 30 cycles of 94°C for 30 seconds, 44°C for 30 seconds, 68°C for 30 seconds and final extension at 68°C for 5 minutes. 10 μl of PCR products were stained with Ethidium bromide solution and visualized under a UV light on 2% agarose gels run at 100 volts for 25 to 30 minutes.

### 2.9. Data analysis

The data were entered and analysed using SPSS 17.0 statistical software (SPSS Inc. USA). Socioeconomic index scores was allotted to each child based on the occupation and educational attainment of the parents as described by Oyedeji ^109^. Categorical variables were compared using the Chi square test while continuous variables were compared by the student t-test. The main outcome (dependent) variables were the prevalence of asymptomatic malaria using microscopy, RDT and PCR Values are expressed as percentages. Statistical significance level was set at P < 0.05.

### 2.10. Ethics approval

Approval for the study protocol was obtained from the Oyo State Ministry of Health, Ethical Review Committee (Reference Number AD 13/479/258) as well as Nagasaki University Ethical Review Committee. Participation in the study was completely voluntary and written informed consent obtained from the parents/guardians of all the participants. The results of Haemoglobin electrophoresis, haematocrits and blood groups were printed and made available to all participants.

## 3. Results

### 3.1. Characteristics of study participants

Participants comprised 496 (48.5%) males and 527 (51.5%) females. The age ranged from 10 to 19 years with a mean age of 14.6 ± 2.3 years. 49.1% were aged 10 – 14 years. Socio-demographic characteristics of the participants are given in **Table I**. The majority of participants resided in rural areas (n = 768, 75.1%). Mean age, distribution by social class and age groups were not significantly different between males and females. Conversely, a greater percentage of male participants were thinner and more stunted than their female counterparts (**Table I**).

**Table I:** Characteristics of study participants.

| Characteristics | All participants (1023) | Male (496) | Female (527) | p |
| --- | --- | --- | --- | --- |
| Place of residence |  |  |  |  |
| Rural, n (%) | 768 | 361 (27.2) | 407 (22.8) | 0.100 |
| Urban, n (%) | 255 | 135 (72.8) | 120 (77.2) |  |
| Mean age (years) | 14.6 ± 2.3 | 14.6 ± 2.3 | 14.5 ± 2.2 | 0.729 |
| Age group (years) |  |  |  |  |
| 10 – 14 | 502 | 246 (49.0) | 256 (51.0) | 0.744 |
| 15 – 19 | 521 | 250 (48.0) | 271 (52.0) |  |
| Social class |  |  |  |  |
| Low, n (%) | 503 | 258 (10.1) | 245 (8.7) | 0.082 |
| Middle, n (%) | 424 | 188 (37.9) | 236 (44.8) |  |
| High, n (%) | 96 | 50 (52.0) | 46 (46.5) |  |
| BMI-for-age |  |  |  |  |
| Thin, n (%) | 180 | 122 (67.8) | 58 (32.2) | <0.001 |
| Normal weight, n (%) | 841 | 374 (44.5) | 467 (55.5) |  |
| Overweight, n (%) | 2 | 0 (0.0) | 2 (100) |  |
| Height for age |  |  |  |  |
| Stunted | 229 | 160 (69.9) | 69 (30.1) | <0.001 |
| Normal | 780 | 329 (42.2) | 451 (57.8) |  |
| Tall | 14 | 7 (50.0) | 7 (50.0) |  |

### 3.2. Prevalence of asymptomatic malaria by microscopy, RDT and PCR

Blood film examination by microscopy revealed that 92 out of 1023 had malaria parasitaemia, giving a prevalence of asymptomatic malaria by microscopy of 9% among schooling adolescents (**Figure 1A**). 47% of participants were malaria parasite positive by RDT (484 out of 1023. Of the 658 samples randomly selected for PCR analysis, 80% were positive for malaria parasites (**Figure 1A**).

### 3.3. Factors associated with asymptomatic malaria parasite carriage

We examined the factors associated with the carriage of malaria parasites as detected by PCR amongst the 658 participants selected for PCR analysis. We found that residence in a rural, as opposed to an urban area was strongly associated with an increased risk of malaria parasite carriage (OR (95% CI) 5.91 (0.79, 44.29), P <0.001, **Table II**). Younger adolescents (aged 10-14) were more likely to harbour malaria parasites than older adolescents (15-19) (OR (95% CI) 1.55 (1.02, 2.34) P = 0.039). Asymptomatic malaria parasite carriers were more likely to be below average height for their age (OR (95% CI) 1.72 (1.09, 2.71) P = 0.026).

**Table II:**
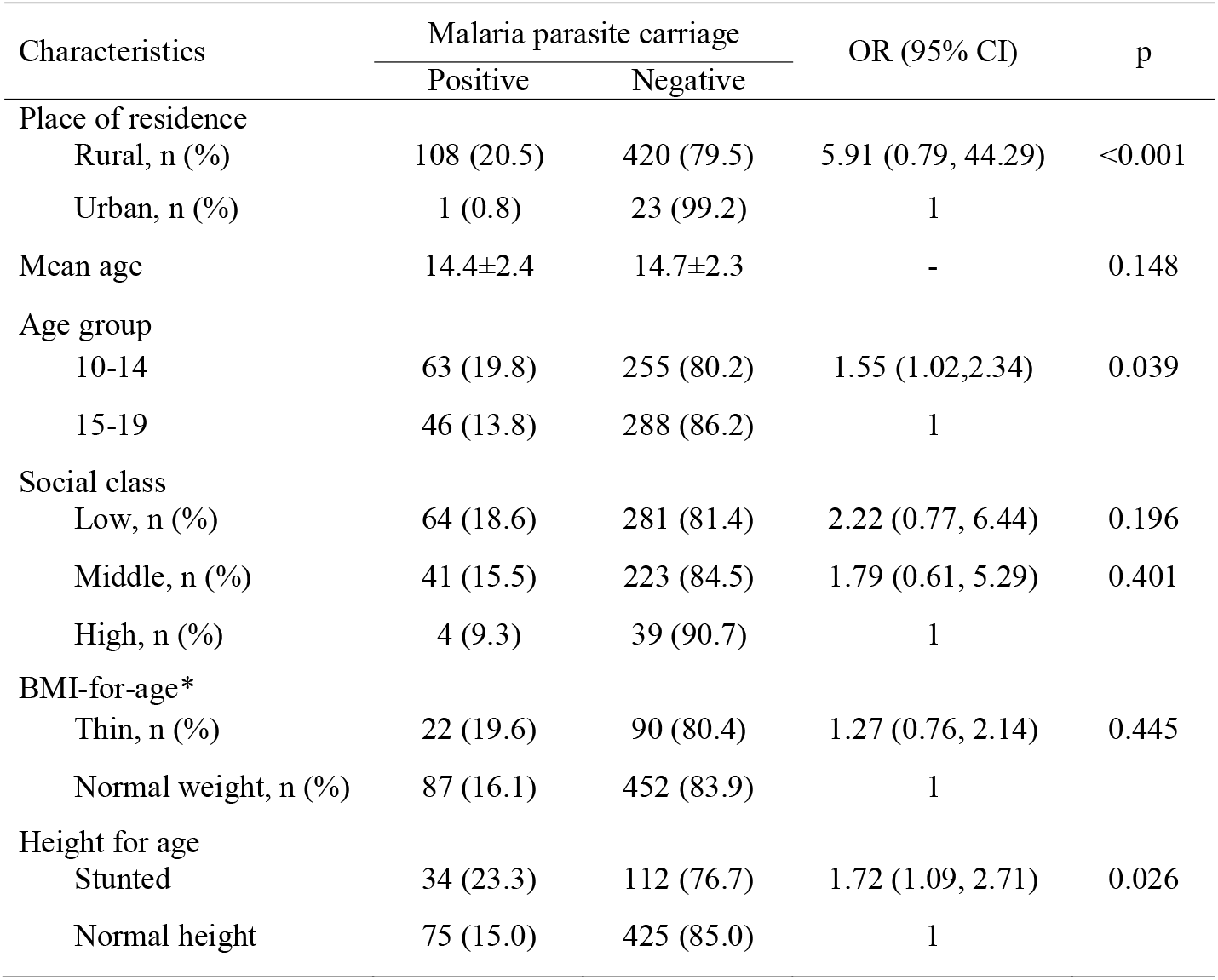
Socio-demographic factors and Asymptomatic *Plasmodium spp.* carriage.

### 3.4. Malaria parasite species prevalence

We determined which species were present in all asymptomatic malaria parasite carriers identified by PCR. Of the 530 PCR positive participants, 455 carried *P. falciparum*, 352 carried *P. malariae*, and 162 carried *P. ovale* parasites (**Figure 1B**). Sixty-two percent of participants carried mixed species infections (**Figure 1C**), with the majority of these involving infection with both *P. falciparum* and *P. malariae* (296 out of 330 mixed infections, 187 of which were double infections of these two species with the remaining 109 being triple infections including *P. ovale* (Figure 1B and D). Plasmodium falciparum plus *P. ovale* double infections accounted for 34 infections. There were 125, 56 and 19 single infections of *P. falciparum, P. malariae* and *P. ovale*, respectively.

### 3.5. Factors associated with species specific asymptomatic malaria parasite carriage

Following PCR determination of the species composition of asymptomatic malaria infections, we determined the risk factors for the carriage of each species in single and mixed infections. Carriage of *P. falciparum* was associated with living in a rural area (OR = 6.24; 95% CI = 4.10, 9.49, P < 0.001), being of low (OR = 3.20; 95% CI = 1.68, 6.11, P < 0.001) or middle (OR = 2.10; 95% CI = 1.09, 4.02, P =0.036) social class and being below normal height (OR = 2.06; 95% CI = 1.31, 3.23, P < 0.01) (**Table III**).

**Table III:** Socio-demographic factors and *Plasmodium falciparum* carriage.

| Characteristics | *P. falciparum* carriage |  | OR (95% CI) | p |
| --- | --- | --- | --- | --- |
|  | Positive | Negative |  |  |
| Place of residence |  |  |  |  |
| Rural, n (%) | 412 (78.0) | 116(22.0) | 6.24 (4.10, 9.49) | <0.001 |
| Urban, n (%) | 45 (36.3) | 79(63.7) | 1 |  |
| Mean age | 14.6±2.2 | 14.7±2.2 | - | 0.628 |
| Age groups (years) |  |  |  |  |
| 10 – 14 | 233 (73.3) | 85 (26.7) | 1.35 (0.96, 1.89) | 0.083 |
| 15 – 19 | 224 (67.1) | 110 (32.9) |  |  |
| Social class |  |  |  |  |
| Low, n (%) | 260 (75.4) | 85(24.6) | 3.20 (1.68, 6.11) | <0.001 |
| Middle, n (%) | 176 (66.7) | 88(33.3) | 2.10 (1.09, 4.02) | 0.036 |
| High, n (%) | 21 (48.8) | 22(51.2) | 1 |  |
| BMI-for-age* |  |  |  |  |
| Thinnest, n (%) | 85 (75.9) | 27 (24.1) | 1.43 (0.89, 2.28) | 0.170 |
| Normal weight, n (%) | 371 (68.8) | 168 (31.2) | 1 |  |
| Height for age |  |  |  |  |
| Stunted | 118 (80.8) | 28 (19.2) | 2.06 (1.31, 3.23) | 0.002 |
| Normal height | 336 (67.2) | 164 (32.8) | 1 |  |

\*Only 2 participants are overweight

Carriage of *P. malariae* was not associated with socio-economic status, but was associated with living in rural areas (OR = 3.88; 95% CI = 2.54, 5.94, P < 0.001), younger age (OR = 1.36; 95% CI = 1.00, 1.86, P = 0.05), being below normal BMI (OR = 1.99; 95% CI = 1.29, 3.07, P < 0.002), and being below normal height (OR = 1.53; 95% CI = 1.05, 2.24, P < 0.034) (**Table IV**).

**Table IV:** Socio-demographic factors and *Plasmodium malariae* carriage.

| Characteristics | *P. malariae* carriage |  | OR (95% CI) | p |
| --- | --- | --- | --- | --- |
|  | Positive | Negative |  |  |
| Place of residence |  |  |  |  |
| Rural, n (%) | 324 (61.4) | 204 (38.6) | 3.88 (2.54, 5.94) | < 0.001 |
| Urban, n (%) | 36 (29.0) | 88 (71.0) | 1 |  |
| Mean age | 14.5±2.1 | 14.8±2.4 | - | 0.130 |
| Age groups (years) |  |  |  |  |
| 10 – 14 | 188 (59.1) | 130 (40.9) | 1.36 (1.00, 1.86) | 0.050 |
| 15 – 19 | 172 (51.5) | 162 (48.5) | 1 |  |
| Social class |  |  |  |  |
| Low, n (%) | 200(58.0) | 145(42.0) | 1.59 (0.84, 3.00) | 0.205 |
| Middle, n (%) | 120(53.0) | 124(47.0) | 1.11 (0.58, 2.13) | 0.874 |
| High, n (%) | 20(46.5) | 23(53.5) | 1 |  |
| BMI-for-age* |  |  |  |  |
| Thin, n (%) | 77 (68.8) | 35 (31.2) | 1.99 (1.29, 3.07) | 0.002 |
| Normal weight, n (%) | 283 (52.5) | 256 (47.5) | 1 |  |
| Height for age |  |  |  |  |
| Stunted | 93 (63.7) | 53 (36.3) | 1.53 (1.05, 2.24) | 0.034 |
| Normal height | 267 (53.4) | 233 (46.6) | 1 |  |

Asymptomatic carriage of *P. ovale* was associated with living in a rural region (OR = 7.46; 95% CI = 3.41, 16.36, P < 0.001) and being of middle (OR = 4.31; 95% CI = 1.49, 12.42, P < 0.007) or low (OR = 3.92; 95% CI = 1.37, 11.27, P < 0.011) socio-economic status (**Table V**).

**Table V:** Socio-demographic factors and *Plasmodium ovale* carriage.

| Characteristics | *P. ovale* carriage |  | OR (95% CI) | p |
| --- | --- | --- | --- | --- |
|  | Positive | Negative |  |  |
| Place of residence |  |  |  |  |
| Rural, n (%) | 163(30.9) | 365(69.1) | 7.46 (3.41, 16.36) | <0.001 |
| Urban, n (%) | 7(5.6) | 117(94.4) | 1 |  |
| Mean age | 14.7±2.1 | 14.6±2.3 | - | 0.824 |
| Age group (years) |  |  |  |  |
| 10 - 14 | 89 (28.0) | 229 (72.0) | 1.21 (0.86, 1.72) | 0.277 |
| 15 – 19 | 81 (24.3) | 253 (75.7) | 1 |  |
| Social class |  |  |  |  |
| Low, n (%) | 99 (28.7) | 246 (71.3) | 3.92 (1.37, 11.27) | 0.011 |
| Middle, n (%) | 87 (25.4) | 197 (74.6) | 4.31 (1.49, 12.42) | 0.007 |
| High, n (%) | 4 (9.3) | 39 (90.7) | 1 |  |
| BMI-for-age* |  |  |  |  |
| Thinnest, n (%) | 29 (25.9) | 83 (74.1) | 0.99 (0.62, 1.57) | 0.952 |
| Normal weight, n (%) | 141 (26.2) | 398 (73.8) | 1 |  |
| Height for age |  |  |  |  |
| Stunted | 47 (32.2) | 99 (67.8) | 1.46 (0.97, 2.18) | 0.084 |
| Normal height | 123 (24.6) | 377 (75.4) |  |  |

**Table VI:** Socio-demographic factors and mixed *Plasmodium falciparum* and *Plasmodium malariae* carriage.

| Characteristics | *P. falciparum* and *P. malariae*<br>species carriage |  | OR (95% CI) | p |
| --- | --- | --- | --- | --- |
|  | Positive | Negative |  |  |
| Place of residence |  |  |  |  |
| Rural, n (%) | 281 (53.2) | 247 (46.8) | 8.94 (5.0,15.91) | <0.001 |
| Urban, n (%) | 14(11.3) | 110(88.7) | 1 |  |
| Mean age | 14.5±2. 1 | 14.7±2. 3 |  | 0.212 |
| Age group |  |  |  |  |
| 10-14 | 158 (49.7) | 160 (50.3) | 1.42 (1.04, 1.94) | 0.032 |
| 15-19 | 137 (41.0) | 197 (59.0) | 1 |  |
| Social class |  |  |  |  |
| Low, n (%) | 168 (48.7) | 177 (51.3) | 1.97 (1.00, 3.85) | 0.052 |
| Middle, n (%) | 113 (42.8) | 151 (57.2) | 1.55 (0.78, 3.07) | 0.272 |
| High, n (%) | 14 (32.6) | 29 (67.4) | 1 |  |
| BMI-for-age* |  |  |  |  |
| Thin, n (%) | 65 (58.0) | 47 (42.0) | 1.86 (1.23, 2.81) | 0.004 |
| Normal weight, n (%) | 230 (42.7) | 309 (57.3) | 1 |  |
| Height for age |  |  |  |  |
| Stunted | 87 (59.6) | 59 (40.7) | 2.07 (1.42, 3.01) | <0.001 |
| Normal height | 208 (41.6) | 292 (58.4) | 1 |  |

Carriage of both *P. falciparum* and *P. malariae* in mixed infections was associated with younger adolescents (OR = 1.42; 95% CI = 1.04, 1.94, P =0.032), living in rural as opposed to urban areas (OR = 8.94; 95% CI = 5.0, 15.91, P < 0.001), and with below average weight and height (OR = 1.86, 95% CI = 1.23, 2.81, P = 0.004; OR = 2.07, 95% CI = 1.42, 3.01, P < 0.001), respectively (**Table VII**).

### 3.6. Comparison of species composition in asymptomatic and symptomatic infections

In order to investigate whether the species composition we observed in asymptomatic adolescents was consistent with symptomatic infections in the same age group, we collected a further 80 samples from a single school in Iroko, and 90 samples from symptomatic malaria patients from a nearby hospital. There were no significant differences in the mean age of participants/patients from these two collections (symptomatic average age 15 years, asymptomatic average age 13 years 8 months, Student’s two-tailed t-test, P > 0.05), and both collections occurred in parallel. We found a similar species composition in the asymptomatic malaria parasite carriers as in the collection from the previous year (**Figure 1C, Figure 2A**), with 64% of infected samples containing mixed infections, the majority of which were *P. falciparum* and *P. malariae* (**Figure 2A**). In stark contrast, 91% of the symptomatic infections were single infections of *P. falciparum*, with only 8 patients out of 90 harbouring mixed infections with *P. malariae* (n= 4) and *P. ovale* (4) (**Figure 2**).

**Figure 2.**
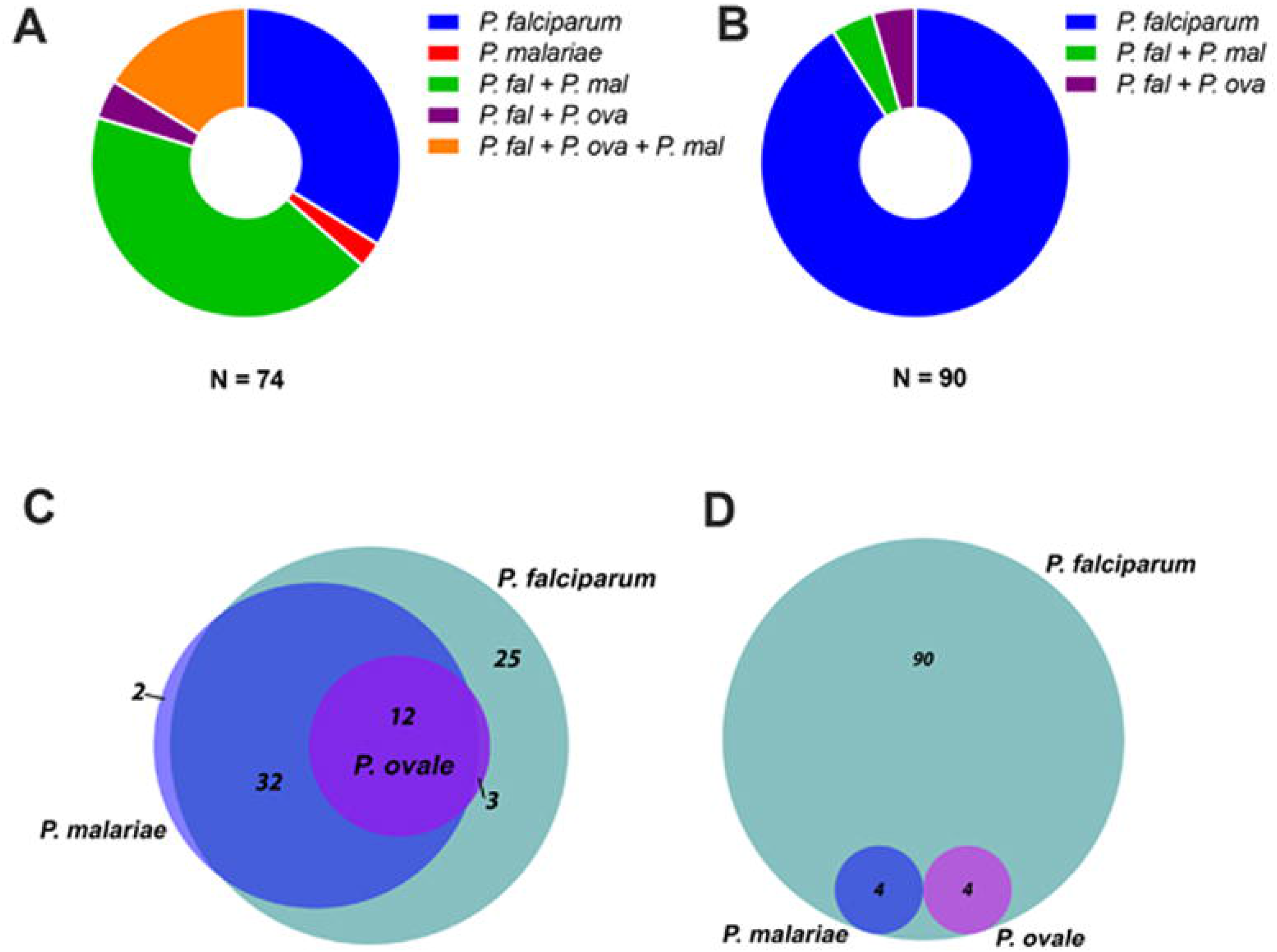
Species composition of 74 asymptomatic and 90 symptomatic infections collected from age-matched individuals in Akinyele region, Ibadan in 2017. (A) Species composition of asymptomatic malaria positive blood samples. (B) Species composition of symptomatic malaria positive blood samples. (C) Prevalence of each of three malaria parasite species assayed (*Plasmodium falciparum, Plasmodium malariae* and *Plasmodium ovale*) in asymptomatic malaria positive blood samples. (D) Prevalence of each of three malaria parasite species assayed in symptomatic malaria positive blood samples.

### 3.7. Multiplicity of Infection of *P. falciparum* clones in single and mixed asymptomatic and symptomatic infections

In order to assess whether symptomatic malaria was caused by the overgrowth of a single clone of *P. falciparum*, we measured the multiplicity of infection (MOI) of *P. falciparum* in symptomatic and asymptomatic infections by typing the polymorphic msp1 gene. We also compared the number of *P. falciparum* msp1 genotypes in asymptomatic *P. falciparum* and *P. malariae* mixed infections with asymptomatic *P. falciparum* single infections.

Symptomatic *P. falciparum* single species infections contained an average of 3.22 msp1 genotypes, compared to 5.75 in asymptomatic single *P. falciparum* infections (Student’s 2-tailed t-test, P <0.01) and 7.18 in asymptomatic mixed species infections of *P. falciparum* and *P. malariae* (Student’s 2-tailed t-test, P <0.01) (**Figure 3A**)

**Figure 3.**
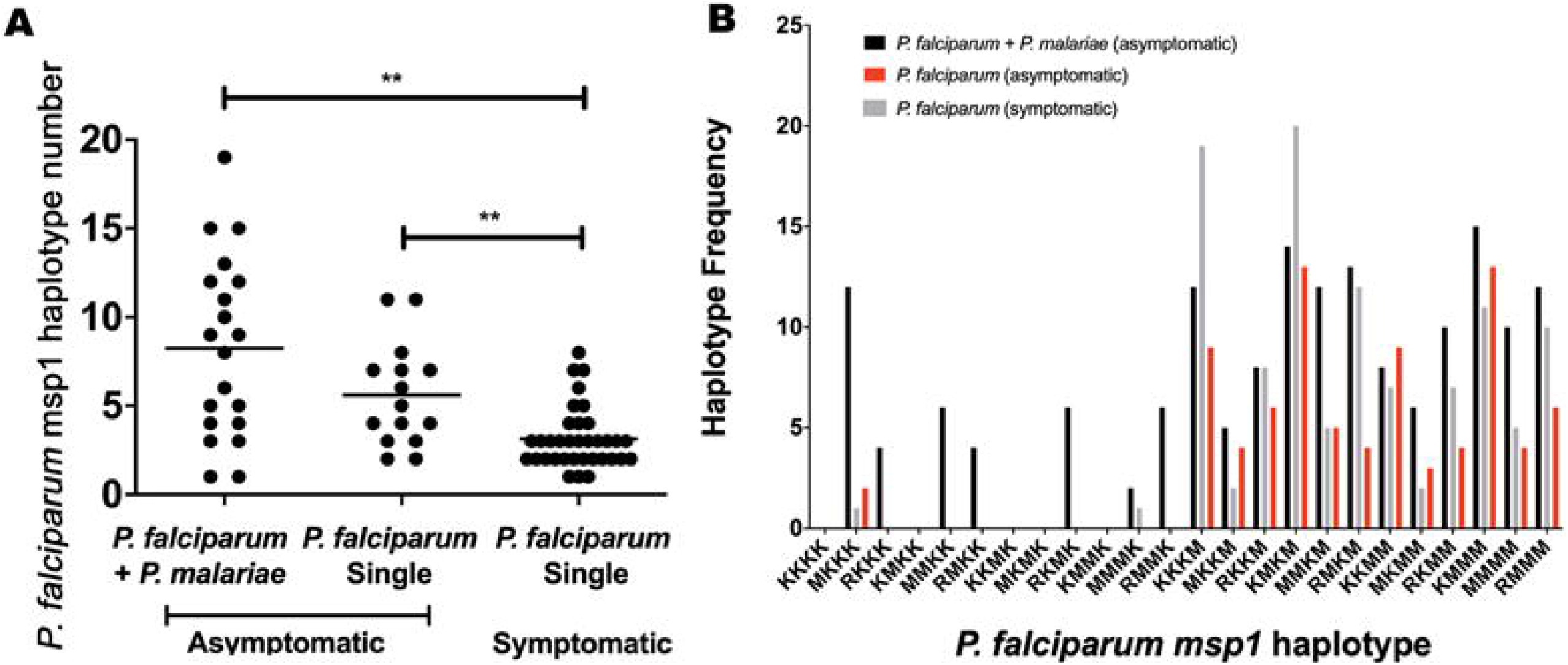
Multiplicity of Infection as determined by haplotyping of the Plasmodium falciparum msp1 gene in symptomatic, asymptomatic single species and asymptomatic mixed *P. falciparum* and Plasmodium malariae infections. (A) Numbers of distinct *P. falciparum* msp1 haplotypes per infection in symptomatic and asymptomatic infections. Horizontal black bars show means (** = P < 0.01, Student’s two-tailed t-tests). (B) Frequency of haplotypes of *P. falciparum* msp1 based on blocks 2, 4a, 4b and 6.

We then asked if the distribution of particular *P. falciparum* msp1 genotypes was common to symptomatic, asymptomatic, and mixed species asymptomatic infections. Several msp1 haplotypes (RKKK, MMKK, RKMK and RMMK) were only found in mixed infections with *P. malariae*, whilst two haplotypes (KKKM and KMKM) were more commonly observed in symptomatic compared to asymptomatic infections (**Figure 3B**)

## 4. Discussion

Nigeria suffers the world’s largest malaria burden, with approximately 51 million cases and 207,000 deaths annually (CDC, 2020). *Plasmodium falciparum* accounts for more than 80% of infections in almost all epidemiological studies on malaria in Nigeria to date(Amodu et al., 2008; Anorlu et al., 2001; Kotila et al., 2007), however, the vast majority of previous studies were conducted on symptomatic infections. Furthermore, most previous studies did not employ molecular analyses such as PCR for malaria parasite species diagnosis. Relatively few surveys have been conducted on the malaria parasite species infecting asymptomatic parasite carriers. Ibekwe et al (2009) surveyed 200 apparently healthy first year university students in the eastern part of Nigeria, mostly aged > 18 years, and found parasitaemia in as high as 80% of the group, with *P. falciparum* accounting for 84%, *P. malariae* 9%, P. vivax 5% and *P. ovale* 2.5%, although only microscopy was used in this study (Ibekwe et al., 2009). However, in another microscopy-based survey, Noland et al (2014) described *P. malariae* prevalence to be as high as 32% amongst symptomatic children less than 10 years old in the South Eastern part of the country (Noland et al., 2014).

Few reports are available on the study of asymptomatic malaria caused by species other than *P. falciparum*. However, like asymptomatic *P. falciparum*, asymptomatic *Plasmodium vivax* malaria has been reported in a range of endemic settings. For example, the low transmission setting of Temotu Province, Solomon Islands and the highly endemic malaria area of Rio Negro in the Amazon State, Brazil both report significant presence of asymptomatic P. vivax (Harris et al., 2010; Suarez-Mutis et al., 2007). Another Amazonian study reported that the prevalence of “symptomless” *P. falciparum* and P. vivax malaria infections are 4–5 times higher than symptomatic infections, with a significant positive correlation of symptomless malaria with older age groups (Alves et al., 2002).

Though rare, cases of *P. malariae* and *P. ovale* asymptomatic infection have also reported. A case report of transfusion-transmitted *P. malariae* infection from an asymptomatic donor reported by Scuracchio et al (2011) suggests that *P. malariae* can be harboured asymptomatically (Scuracchio et al., 2011). Supporting this, another study also reported a case of malaria in a Grecian woman due to *P. malariae* whose illness was reactivated after decades of latency (Vinetz et al., 1998). Infection with *P. ovale* can also be asymptomatic (Rojo-Marcos et al., 2011). However, these studies are mostly case reports and extensive longitudinal studies are lacking.

We found an unexpectedly high prevalence of *P. malariae* in asymptomatic schooling adolescents (66% of malaria parasite positive individuals). This prevalence is much higher than previously reported *P. malariae* prevalences in Africa. This could due be due to numerous factors. Firstly, the present survey was conducted in asymptomatic individuals, whereas the majority of previous parasite species surveys have sampled symptomatic infections only.

Secondly, the PCR assay used here targets the mitochondrial genome-located CoxIII gene (Isozumi et al., 2015). The most commonly used PCR assay in previous surveys utilises the 18S sRNA gene (Snounou et al., 1993). The mitochondrial genome is present at much higher copy numbers per parasite than the 18S rRNA gene (20-150 copies compared to 4-8 copies), and so offers increased sensitivity to PCR assays targeting it (Lloyd et al., 2018). We propose that PCR assays targeting the mitochondrial genome offer increased sensitivity over those targeting lower copy-number nuclear genes, and are preferable for molecular epidemiological studies involving asymptomatic malaria parasite carriers.

Thirdly, the prevalences of malaria parasite species vary across space and time, making meaningful comparisons between surveys carried out at different locations and at different times difficult. Multi-centre longitudinal studies should be conducted across Africa in order to gauge the true prevalence of the various species of malaria parasites that infect humans on the continent.

We found some interesting differences regarding the risk factors associated with asymptomatic carriage of the three malaria parasite species assayed here. The risk of carriage of all three species was greater in the rural area compared to the urban areas, presumably due to the greater entomological inoculation rates encountered in the countryside. However, whilst *P. falciparum* and *P. ovale* were both linked to lower socio-economic status, *P. malariae* was not, infecting individuals of all social strata indiscriminately. Furthermore, *P. falciparum* and *P. ovale* carriage was associated with below average BMI, whereas *P. malariae* was not. These observations are difficult to explain, but may point towards differing interactions with host nutritional status between the parasite species.

The prevalence of mixed *P. malariae* and *P. falciparum* co-infections was much higher in asymptomatic compared to symptomatic parasite carriers, the latter of which were almost exclusively infected with single species *P. falciparum*. Interactions between malaria parasite species can have profound impacts on the severity of disease (Tang et al., 2019). This observation is consistent with those of Black et al (1994), who found 27-32% of asymptomatic children under nine years old in the Ivory Coast harboured mixed *P. falciparum* and *P. malariae* infections, while no mixed infections were observed in symptomatic children (Black et al., 1994). In contrast, it was previously shown that *P. malariae* co-infection was a significant risk factor for anaemia in Ghanaian children, increasing its risk 1.6-fold (Ehrhardt et al., 2006).

It is possible that our results may be explained by a protective effect of *P. malariae* co-infection on the severity of *P. falciparum*. It has been suggested that an ongoing *P. malariae* infection may reduce the parasitaemia of a resulting *P. falciparum* infection by up to 50%, due to within-host competition (Mason et al., 1999). There may also be host-immune modulated cross-protection, in which antibodies elicited against one species may be functionally protective against another. There is also the possiblity that *P. malariae* infection may make hosts more susceptible to infection with *P. falciparum*; Domarle et al (1999) reported that they commonly observed *P. malariae* in co-infection with *P. falciparum*. They observed that individuals whose co-infections were cleared with antimalarial drugs succumbed to additional infections of *P. falciparum* more quickly than individuals who were previously infected with *P. falciparum* single species infections (Domarle et al., 1999).

The absence of *P. malariae* co-infection in symptomatic patients may be caused by the overgrowth of one or more clones of *P. falciparum*. This could have lead to the competetive exclusion of *P. malariae* either through within host-competition for host blood cells (Tang et al., 2019), or through the raising of an acute inate immune response in the host that kills off the slower growing species (Molineaux et al., 1980). Supporting this is our observation that the multiplicity of *P. falciparum* infection in symptomatic patients is significantly lower than in asymptomatic parasite carriers.

One further possible explanation for our observation is due to the nature of the nested PCR approach used for species diagnosis. Given that the first round of the PCR uses primers that amplify a portion of the CoxIII gene conserved among *Plasmodium spp*, followed by a species-specific amplification round, it is possible that the assay will predominantly amplify the dominant species in an infection. In the case of symptomatic infections, in which the parasitaemia of *P. falciparum* is high, it is possible that that relatively low levels of *P. malariae* may be missed by this assay.

Complexity of infections, including the number of clones and species have been linked to increased anaemia in asymptomatic Nigerian children (May et al., 2000). In the present study, we found that symptomatic malaria was associated with a lower number of *P. falciparum* clones per infection than asymptomatic infection. *Plasmodium falciparum* multiplicity of infection was highest in mixed species infections with *P. malariae*, and several *P. falciparum* msp1 haplotypes were only observed in co-infections with *P. malariae*. These results are consistent with the hypothesis that overgrowth of particular *P. falciparum* clones leads to the competitive exclusion of other clones, and other species in the infection, and leads to the development of clinical symptoms in the host. Further surveys are required to gather more data regarding whether particular *P. falciparum* strains are more likely to be found in mixed species infections than others.

In conclusion, we found a much higher than expected prevalence of *P. malariae* and *P. ovale* infections in asymptomatic adolescents in Ibadan province, southeast Nigeria. Mixed species infections were far more common in asymptomatic compared to symptomatic infections. In order to determine if this pattern is specific to this area and time, or whether these results represent the broad trend of *P. malariae* and *P. ovale* epidemiology across Africa, further multi-centre longitudinal studies are required, preferably utilising highly sensitive molecular epidemiological assays.

## Data Availability

All data associated with this work is presented in the manuscript.

## Acknowledgements

R.C. is supported by Japanese Society for the Promotion of Science (JSPS), Japan Grant-in-Aid for Scientific Research Nos. 16K21233 and 19K07526

